# Variants in *MARC1* and *HSD17B13* reduce severity of NAFLD in children, perturb phospholipid metabolism, and suppress fibrotic pathways

**DOI:** 10.1101/2020.06.05.20120956

**Authors:** Christian A. Hudert, Anna Alisi, Quentin M. Anstee, Annalisa Crudele, Laura G. Draijer, EU-PNAFLD investigators, Samuel Furse, Jan G. Hengstler, Benjamin Jenkins, Kylie Karnebeek, Deirdre A. Kelly, Bart G. Koot, Albert Koulman, David Meierhofer, Stuart G. Snowden, Indra van Mourik, Anita Vreugdenhil, Susanna Wiegand, Jake P. Mann

## Abstract

**Background & aims:** Genome-wide association studies in adults have identified variants in *HSD17B13* and *MARC1* as protective against NAFLD. It is not known if they are similarly protective in children and, more generally, whether the peri-portal inflammation of pediatric NAFLD and lobular inflammation seen in adults share common genetic influences. Therefore, we aimed to: establish if these variants are associated with NAFLD in children, and to investigate the function of these variants in hepatic metabolism using metabolomics.

**Methods:** 960 children (590 with NAFLD, 394 with liver histology) were genotyped for rs72613567T>TA in *HSD17B13*, rs2642438G>A in *MARC1*. Genotype-histology associations were tested using ordinal regression. Untargeted hepatic proteomics and plasma lipidomics were performed in a subset of samples. *In silico* tools were used to model the effect of rs2642438G>A (p.Ala165Thr) on *MARC1*.

**Results:** rs72613567T>TA in *HSD17B13* was associated with lower odds of NAFLD diagnosis (OR 0.7 (95%CI 0.6-0.9) and lower grade of portal inflammation (P<0.001) whilst rs2642438G>A in *MARC1* was associated with lower grade of hepatic steatosis (P=0.02). Proteomics found reduced expression of HSD17B13 in carriers of the protective allele, whereas MARC1 levels were not affected by genotype. Both variants showed downregulation of hepatic fibrotic pathways, upregulation of retinol metabolism and perturbation of phospholipid species. Modelling suggests that p.Ala165Thr would disrupt the stability and metal-binding of *MARC1*.

**Conclusions:** There are shared genetic mechanisms between pediatric and adult NAFLD, despite their differences in histology. *MARC1* and *HSD17B13* are involved in phospholipid metabolism and suppress fibrosis in NAFLD.

## Introduction

Understanding genetic variants associated with human chronic disease phenotypes has yielded insights into multifactorial pathogenesis^1^. This is of particular importance in non-alcoholic fatty liver disease (NAFLD) as it is a common condition with potential to progress to end-stage liver disease and hepatocellular carcinoma^2,3^, yet has no licensed therapies^4^.

Population level genome- and exome-wide association studies have identified several common variants implicated in the severity of NAFLD^5^. p.Ile148Met in patatin-like phospholipase domain-containing protein 3 (*PNPLA3*) is the variant most strongly associated with increased severity of NAFLD^6,7^ and has been well studied, leading to its identification as a lipid droplet-binding protein that influences the recruitment of hydrolysing enzymes^8,9^. p.Glu167Lys in transmembrane 6 superfamily 2 (*TM6SF2*) is also well-established as a harmful variant that impairs hepatic lipid metabolism and increases risk of all stages of NAFLD^10,11^.

More recently, strong human genetic evidence has identified two protective variants at genome-wide significance: rs72613567T>TA in hydroxysteroid 17-beta dehydrogenase 13 (HSD17B13)^12^ and p.Ala165Thr in mitochondrial amidoxime reducing component 1 (*MARC1*, rs2642438G>A)^13^. *HSD17B13* has also been implicated in altered lipid metabolism, including recently in regulation of phospholipids^14^ and retinol^15^. However the function of *MARC1* in hepatic lipid metabolism is largely unknown, though it clearly has drug detoxifying activity^16,17^ and has been recently implicated in phospholipid metabolism^18^.

Histological validation of genetic variants is challenging due to the comparatively small numbers who undergo liver biopsy for NAFLD. This is even more so the case in pediatric NAFLD, therefore analyses studies are candidate gene studies in relatively small cohorts^19,20^. One genome-wide association study has been conducted in Hispanic boys with biopsy-proven NAFLD, which yielded several suggestive loci^21^. It is still unclear precisely how closely the genetics of adult and pediatric NAFLD overlap.

Pediatric NAFLD is common (7.6% in the general population^22^) and shows a predominance of peri-portal inflammation and zone 1 steatosis, particularly in younger male patients^23,24^. Whilst both are positively associated with insulin resistance and other features of the metabolic syndrome^25^, it is not clear whether pediatric peri-portal inflammation in NAFLD is simply a childhood manifestation of ‘adult’ NASH (with lobular inflammation and ballooning) or a different pathophysiological entity.

Therefore, we sought to address whether the protective variants in *HSD17B13* and *MARC1* identified at a population-level in adults can be replicated in children with NAFLD, also providing histological validation of p.Ala165Thr in *MARC1*. In this study we used liver tissue proteomics and plasma lipidomics to gain insight into the impact of these variants on hepatic metabolism in children with NAFLD.

## Methods

### Participants

Two groups of participants were included in this cross-sectional study: cases (children with NAFLD) and controls (without NAFLD). A subset of children with NAFLD had undergone liver biopsy for clinical indications. All participants (or their parents) gave written informed consent and were recruited between 2014-2019.

Participants were recruited from six sites. Children with NAFLD were recruited from hepatology & gastroenterology clinics in Cambridge & Birmingham (UK) as part of the European Paediatric NAFLD Registry (EU-PNAFLD, Clintrials.gov NCT:04190849)^26^, which was approved by the East Midlands - Nottingham 2 Research Ethics Committee (17/EM/0084).

Obese controls and NAFLD cases were recruited from pediatric obesity clinics at Maastricht University Medical Centre (under ethical approval METC 13-4-130) and Amsterdam University Medical Centre (under ethical approval MEC 2017_306 and MEC 07/141). Obese controls and NAFLD cases were recruited from Bambino Gesu hospital (under ethical approval for EU-PNAFLD and local Ethics Review Board, protocol number 1774_OPBG_2019). Children were referred to these clinics due to obesity and were then subsequently investigated for comorbidities, including NAFLD. Children identified to have NAFLD were recruited as cases and those with obesity but no evidence of liver disease (as described below) were included in this study as obese controls.

In addition, NAFLD cases were recruited from the Berlin Cohort^27^ at the pediatric obesity outpatient clinic and pediatric gastroenterology outpatient clinic of Charité (under ethical approval of the local institutional review board EA2/049/14).

Recruitment of controls from the same clinics (and referral populations) as cases aimed to reduce the bias of case-control comparisons. Recruitment from multiple different hospitals aimed to reduce bias associated with a sample from a single hospital population. As an exploratory analysis, we utilised data from the maximum number of available participants therefore no formal sample size calculation was performed.

### Inclusion and exclusion criteria

All participants were 5-18 years old at the time of inclusion.

Cases (n=590): All cases were identified with abnormal biochemistry (alanine aminotransferase (ALT) >44U/l for girls or >50 U/l for boys and/or aspartate aminotransferase (AST) >50 U/l^28^) and/or radiological evidence of steatosis (ultrasound, magnetic resonance imaging, or controlled attenuation parameter ≥240). Imaging was available for 571 patients (97%). Liver biopsy with histological characterization of NAFLD was available for 394 (67%) patients.

Controls (n=412): NAFLD was excluded by absence of hepatic steatosis upon radiologic examination (ultrasound, magnetic resonance imaging, or controlled attenuation parameter <240), or where not available (n=177, 43%) with normal aminotransferases.

Exclusion criteria were: age <5 or >18 years, any other liver disease (assessment for secondary causes including: alpha-1-antitrypsin deficiency, celiac disease, autoimmune hepatitis, viral hepatitis A, B and C, active cytomegalovirus or Epstein-Barr virus infection, thyroid disorders, and Wilson’s disease), severe underlying chronic disease (e.g., cardiopulmonary or autoimmune disease), alcohol consumption >20 g of alcohol per day, and pregnancy.

Children with incomplete data, inadequate genotyping quality or unclear genotyping call were also excluded. Thirty-six patients initially recruited were excluded, 32 were under 5 or over 18 years old, 2 with unclear diagnosis, and 4 without adequate data.

### Clinical and laboratory investigations

In all cases and obese controls included in the study, anthropometric measures (height, weight) were taken, laboratory analysis including a hepatic panel and complete blood count was performed, and fasted state metabolic parameters were assessed by using standardized procedures. The homeostatic model assessment of insulin resistance (HOMA-IR) was derived using fasting insulin (μU/L) x fasting glucose (nmol/L) / 22.5. Obesity was defined as body mass index (BMI) z-score >2.

### Genotyping

DNA was extracted from whole blood (using Qiagen DNeasy kit #69504). Participants were genotyped by quantitative polymerase chain reaction using TaqMan assays (ThermoFisher #4351379): rs738409C>G in *PNPLA3*, rs58542926C>T in transmembrane 6 superfamily 2 (*TM6SF2*), rs2642438G>A in *MARC1*, and rs72613567T>TA in *HSD17B13* (using the custom sequence from Pirola et al.^29^). These variants were selected due to their evidence as genome-wide risk factors for NAFLD and cirrhosis in adults^6,7,10,12,13,30,31^. Data on variants in *HSD17B13* and *MARC1* were available for 960 participants whilst data on *PNPLA3* variants were available for 490 participants and data on *TM6SF2* variants were available for 489 participants.

All variants were within Hardy-Weinberg equilibrium in the control groups using Chi-squared test: rs58542926C>T in *TM6SF2, P* = 0.08; rs2642438G>A in *MARC1, P* = 0.21; and rs72613567T>TA in *HSD17B13, P* = 0.56, rs738409C>G in *PNPLA3, P* = 0.53.

### Liver biopsies

Liver biopsies were evaluated and scored by experienced pathologists of the respective centers. Staging and grading was performed according to the histological scoring system for non-alcoholic fatty liver disease by the Nonalcoholic Steatohepatitis Clinical Research Network (NASH-CRN)^32^. Briefly, grading included the scoring of steatosis (0= <5%; 1= 5-33%; 2= 34-66%; 3= ≥67%), lobular inflammation (0= 0/200x field; 1 = <2 foci/200x field; 2= 2-4 foci/200x field; 3= >4 foci/ 200x field), and hepatocellular ballooning (0= none; 1= few; 2= many/prominent). Portal inflammation was evaluated according to Brunt et al.^33^ (0= none; 1= mild; 2 = moderate to severe). Staging of fibrosis was performed using NASH-CRN criteria (0= no fibrosis; 1= zone 3 perisinusoidal only or portal/periportal without bridging only; 2= zone 3 perisinusoidal + portal/periportal; 3= bridging fibrosis; 4= cirrhosis).

### Hepatic proteomic analysis

Hepatic tissue proteomics was performed in a subset of 70 patients, as previously described^27^. Liver biopsy specimens were extracted under denaturing conditions and digested by trypsin for subsequent analysis by mass spectrometry^34^. The software tools MaxQuant^35^ and gene set enrichment analysis (GSEA)^36^ were used for peptide identification and pathway analyses, respectively (see Supplementary Methods for details).

### *In silico* analysis of MARC1 p.Ala165Thr

Our hepatic proteomics data suggested that this *MARC1* variant did not alter expression of MARC1 protein, unlike for the studied *HSD17B13* variant. Therefore, to further provide some insight into the impact of rs2642438G>A (p.Ala165Thr) in *MARC1* we used a range of bioinformatics tools to perform an in silico analysis of the variant. UniProt^37^ was searched for isoforms of *MARC1* in other species and sequences aligned. Four tools were chosen to be used for prediction of the impact of the missense variant based on recommendations of accuracy from the range of tools available^38,39^: SNPs&GO^40^, PANTHER^41^, Align-GVGD^42^, and MutPred2^43^. For a structural analysis, we used the available crystal structure of MARC1^44^ and three tools for calculating the effect of p.Ala165Thr on overall protein stability: I-Mutant3.0^45^, DUET^46^, and CUPSAT^47^. Data on *in silico* saturation mutagenesis of MARC1 were available from EVmutation. An annotated protein model was generated using UCSF Chimera^48^.

### Plasma lipidomics analysis

Plasma lipidomics was performed in a subset of 129 children with NAFLD who had undergone liver biopsy. The methods have been described in detail elsewhere^49^, but in brief, lipids, triglycerides and sterols were extracted together from fasting plasma using a high throughput technique followed by direct infusion mass spectrometry (DI-MS)^50^.

*Data acquisition--Samples* were infused into an Exactive Orbitrap (Thermo, Hemel Hampstead, UK), using a Triversa Nanomate (Advion, Ithaca, NY, USA). Positive ionisation mode was achieved at a 1·2 kV. The Exactive started acquiring data 20 s after sample aspiration began. The Exactive acquired data with a scan rate of 1 Hz (resulting in a mass resolution of 65,000 full width at half-maximum (fwhm) at 400 m/z). After 72 s of acquisition in positive mode the Nanomate and the Exactive switched over to negative mode, decreasing the voltage to –1·5 kV. The spray was maintained for another 66 s, after which Collision-Induced Dissociation commenced, with a mass window of 50–1000 Da, and was stopped after another 66 s. The sample plate was kept at 10 °C throughout the data acquisition.

*Data processing--Raw* high-resolution mass-spectrometry data were converted to mzXML files and then processed using XCMS (www.bioconductor.org) and Peakpicker v 2.0 (an in-house R script^51^). Lists of known species (by *m/z*) were used for both positive ion and negative ion mode (~8·5k species). Identification of species was compared against validated lists of species used by this group^51-55^. Variables whose mass deviated by more than 9 ppm from the expected value, had a signal-to-noise ratio of <3 and had signals for fewer than 50% of samples were discarded.

Signals were then divided by the sum of signals for that sample and expressed per mille (‰). Zero values were interpreted as not measured.

### Statistical analyses

Testing for normal distribution was performed for all variables using the Shapiro-Wilk method. Frequencies and percentages are presented for clinical (categorial) and histological (ordinal) characteristics. Medians and quartiles of continuous anthropometric and laboratory parameters were calculated for the total study population as well as for subgroups (stratification for the presence of NAFLD, variant genotype within cases, patients with proteomic or lipidomic profiles). To evaluate differences in distribution between cases and controls as well as within genotypes of variants we used Kruskal-Wallis test for continuous nonparametric values (age, BMI-z score). Chi-squared tests were applied to categorical and ordinal variables (gender and presence of obesity). For all continuous laboratory values, linear regression models with correction for age and gender were used.

Hardy-Weinberg equilibrium was tested using a chi-squared test with one degree of freedom for all genetic variants. Case-control studies between the different genotypes were analyzed using chi-squared tests and logistic regression models for distinct genetic modes of inheritance: no particular genetic model (genotypes), additive (trend), dominant, recessive or multiplicative (alleles). Additive (or linear mixed) genetic models were used in the identification of these variants as significant risk loci^12,13^ and therefore we have primarily used the additive model in all analyses. Effects of variants were calculated using Wald tests, odds ratios (OR) and 95% confidence intervals (95% CIs). False discovery rate-adjusted Q-values were calculated using the Benjamini-Hochberg procedure to adjust for multiple testing. Associations between genotypes and histological features were tested using univariate ordinal regression or multivariate ordinal regression models with correction for age, sex, BMI z-score and HOMA-IR. Dichotomous histological associations were tested by binary logistic regression with correction for age and sex. OR and 95% CIs were calculated. Genotype was coded by number of ‘protective’ alleles: *HSD17B13* rs72613567 T/T=0, T/TA=1, TA/TA=2; and *MARC1* rs2642438 G/G=0, G/A=1, A/A=2. Proteomic studies: see Supplementary Methods.

Lipidomic studies: relative abundances for each lipid were logarithmically transformed and standardised (to mean = 0, standard deviation = 1). Missing values were ignored and no imputation was used. Linear regression analyses adjusted for sex and age were run to test for associations between lipid species and genotype. The beta regression coefficients were then plotted against length of carbon chain and number of double-bonds. Due to high correlation between lipid species, the critical p-value for significance was defined by 0.05√n, where n is the number of identified species from each analysis method.

Statistical analysis was performed in SPSS (IBM Corp. Released 2017. IBM SPSS Statistics for Windows, Version 25.0. Armonk, NY: IBM Corp). Further regression analysis was performed in Stata v16.1 (StataCorp) and random forest analysis was performed using R 3.6.1^56^, in addition, graphs were produced in GraphPad v8.0 for Mac, GraphPad Software, La Jolla California, USA.

## Results

### Characteristics of the study population

One thousand two children (590 with NAFLD and 412 controls) were included in the study. Children with NAFLD were older, more likely to be male, more insulin resistant and dyslipidemic despite being less obese (Table 1). They exhibited higher liver transaminases, with median ALT being elevated more than 2 times as compared to the control group.

**Table 1.** Clinical and Laboratory Characteristics. Data represent frequencies (%) or median (interquartile range) as appropriate. For clinical characteristics, P-values were calculated using Mann-Whitney U test for continuous traits and Chi-square test for categorical traits. For plasma markers, P-values were calculated using linear regression with correction for age, sex. For genotypes, P-values were calculated using binary logistic regression with correction for age and sex (Wald test). FDR correction (Q value) for multiple comparisons was calculated using the Benjamini and Hochberg method. BMI, body mass index; ALT, alanine aminotransferase; AST, aspartate aminotransferase; GGT, gamma glutamyl transferase; LDL, low density lipoprotein; HDL, high density lipoprotein; HOMA, homeostatic model assessment of insulin resistance.

| Variable | Control (N = 412) | NAFLD (N = 590) | P value | Q value |
| --- | --- | --- | --- | --- |
| Age (years) | 12.3 (10.0 – 14.7) | 13.0 (11.0 – 15.4) | **$7.6 \times 10^{-5}$** | **$1.2 \times 10^{-4}$** |
| Male sex, n (%) | 181 (43.9) | 386 (66.6) | **$1.1 \times 10^{-8}$** | **$2.9 \times 10^{-8}$** |
| BMI z-score | 3.1 (2.6 – 3.5) | 2.4 (1.8 – 3.0) | **$1.6 \times 10^{-25}$** | **$8.5 \times 10^{-25}$** |
| Obesity, n (%) | 325 (78.9) | 387 (66.5) | **$2.2 \times 10^{-5}$** | **$3.9 \times 10^{-5}$** |
| ALT (U/l) | 21 (17 – 28) | 47 (31 – 78) | **$1.4 \times 10^{-37}$** | **$2.2 \times 10^{-36}$** |
| AST (U/l) | 25 (20 – 30) | 38 (28 – 54) | **$2.0 \times 10^{-29}$** | **$1.6 \times 10^{-28}$** |
| GGT (U/l) | 16 (13 – 20) | 23 (16 – 36) | **$1.5 \times 10^{-14}$** | **$6.0 \times 10^{-14}$** |
| Cholesterol (mg/dl) | 158 (139 – 178) | 160 (139 – 185) | 0.531 | 0.571 |
| LDL (mg/dl) | 93 (75 – 112) | 97 (81 – 112) | 0.488 | 0.571 |
| HDL (mg/dl) | 46 (39 – 54) | 43 (38 – 50) | **$3.0 \times 10^{-7}$** | **$6.9 \times 10^{-7}$** |
| Triglycerides (mg/dl) | 81 (57 – 108) | 98 (71 – 141) | **$4.0 \times 10^{-6}$** | **$8.0 \times 10^{-6}$** |
| HOMA | 2.6 (1.7 – 4.0) | 3.7 (2.5 – 5.6) | **$2.8 \times 10^{-10}$** | **$9.0 \times 10^{-10}$** |
| HSD17B13 genotype, n (%) |  |  |  |  |
| TT / TTA / TATA | 206 (55.7)/143 (38.6)/21 (5.7) | 394 (66.8)/168 (28.5)/28 (4.7) | **0.002** | **0.003** |
| MARC1 genotype, n (%) |  |  |  |  |
| GG / GA / AA | 199 (53.8)/151 (40.8)/20 (5.4) | 324 (54.9)/227 (38.5)/39 (6.6) | 0.889 | 0.889 |
| PNPLA3 genotype, n (%) |  |  |  |  |
| CC / CG / GG | 48 (49.5)/42 (43.3)/7 (7.2) | 139 (35.4)/172 (43.8)/82 (20.9) | **0.001** | **0.001** |
| TM6SF2 genotype, n (%) |  |  |  |  |
| CC / CT / TT | 84 (87.5)/11 (11.5)/1 (1.0) | 337 (85.8)/52 (13.2)/4 (1.0) | 0.535 | 0.571 |

### Association between variants in *MARC1, HSD17B13, TM6SF2, PNPLA3* and NAFLD

First, we sought to determine whether these variants were associated with diagnosis of NAFLD in children (Figure 1). rs738409C>G in *PNPLA3* was positively associated with diagnosis of NAFLD (odds ratio (OR) 1.79 (95% CI 1.26-2.53). rs72613567T>TA in *HSD17B13* was protective against diagnosis of NAFLD (OR 0.70 (95% CI 0.56-0.87)) though no association with diagnosis of NAFLD was observed with rs2642438G>A in *MARC1* or rs58542926C>T in *TM6SF2* (Supplementary Table 1).

**Figure 1.**
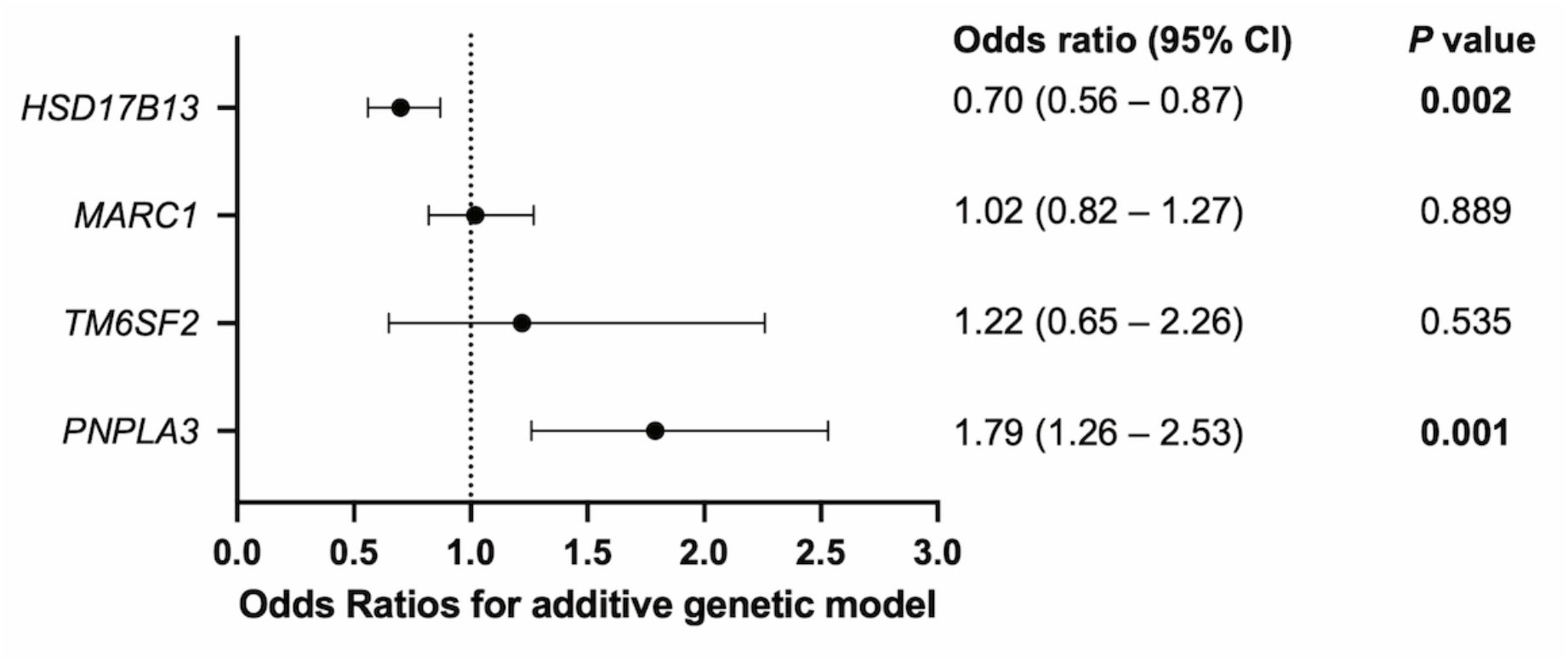
Odds ratios for the presence of NAFLD using an additive genetic model. Data from 590 children with NAFLD (cases) and 370 controls. *P*-values were calculated using logistic regression with correction for age and sex. *‘PNPLA3’* refers to rs738409C>G, *‘MARC1’* refers to rs2642438G>A, and *‘HSD17B13’* refers to rs72613567T>TA and *‘TM6SF2‘* refers to rs58542926C>T.

In children with NAFLD, rs72613567T>TA in *HSD17B13* and rs2642438G>A in *MARC1* were not associated with any anthropometric or biochemical traits (Supplementary Tables 2 & 3).

### Effect of single nucleotide variants on histological severity of NAFLD

Of those with NAFLD, 394 (67%) had undergone liver biopsy (Table 2 & Supplementary Table 4). Participants displayed the whole spectrum of NAFLD from simple steatosis to NASH-associated cirrhosis. 70% had evidence of peri-portal inflammation and 13% had advanced fibrosis (stage 3-4).

**Table 2.** Histologic Findings. Data represent frequencies (%) of histologic findings in the biopsied study population (N = 394).

| Grade / Stage | Steatosis<br>(N= 394) | Portal Inflammation<br>(N = 386) | Lobular Inflammation<br>(N = 394) | Ballooning<br>(N = 388) | Fibrosis<br>(N = 393) |
| --- | --- | --- | --- | --- | --- |
| 0 | - | 117 (30.3) | 77 (19.5) | 121 (31.2) | 90 (22.9) |
| 1 | 90 (22.8) | 226 (58.5) | 202 (51.3) | 161 (41.5) | 152 (38.7) |
| 2 | 187 (47.5) | 43 (11.1) | 115 (29.2) | 106 (27.3) | 99 (25.2) |
| 3 | 117 (29.7) | - | - | - | 50 (12.7) |
| 4 | - | - | - | - | 2 (00.5) |

Consistent with its well-established harmful effect on NAFLD in adults, rs738409C>G in *PNPLA3* was associated with a higher grade of steatosis (P = 3.2×10^-5^), lobular inflammation (P = 0.02, and fibrosis stage (P = 0.02) on multivariable ordinal regression adjusted for age, sex, BMI, and HOMA-IR (Figure 2 & Supplementary Table 5).

**Figure 2.**
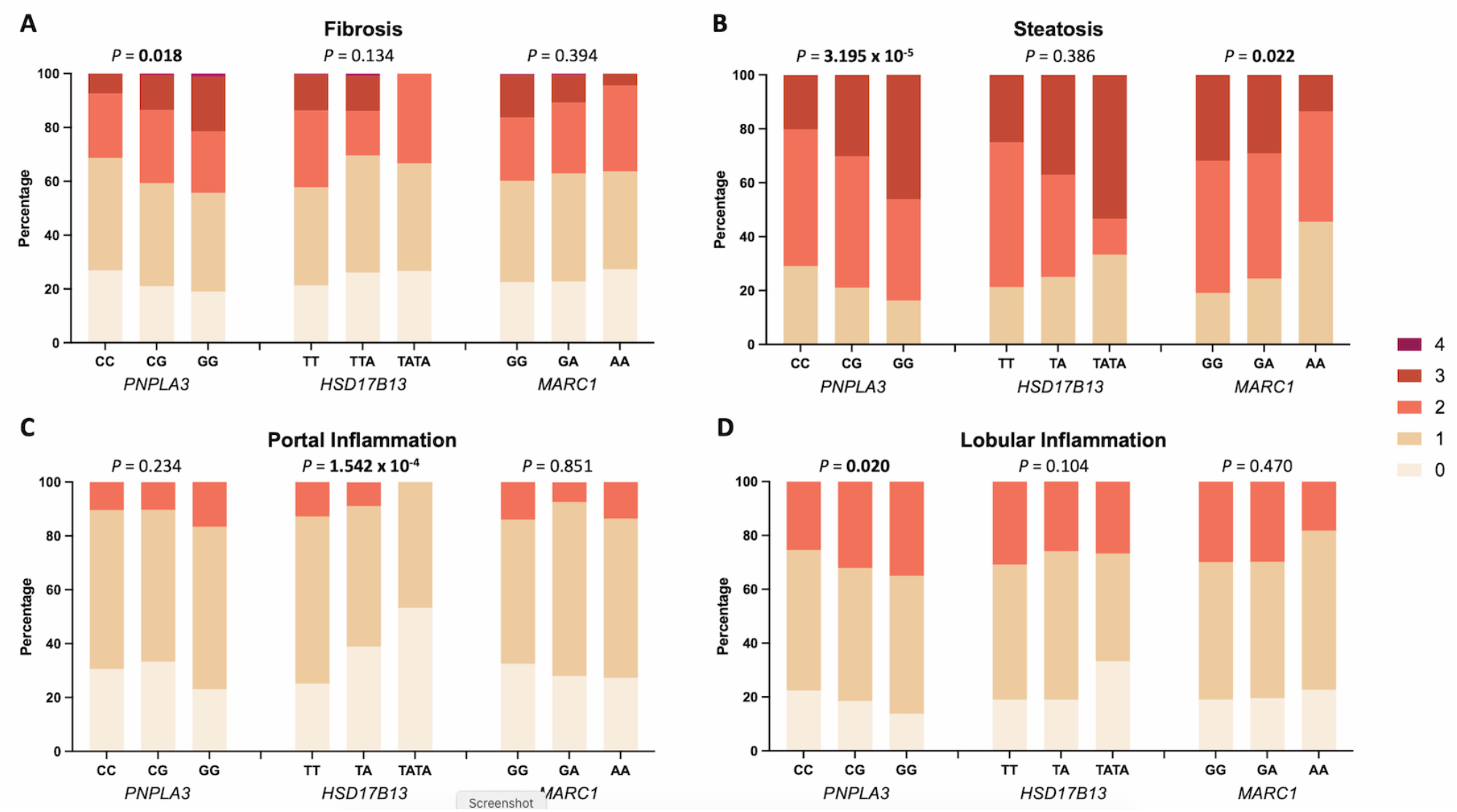
Effect of SNV on histological severity of NAFLD in children. Data from 394 children genotyped for rs738409C>G (*PNPLA3*), rs72613567T>TA (*HSD17B13*) and rs2642438G>A (*MARC1*). Multivariate ordinal regression analysis with correction for age, sex, BMI z-score and HOMA-IR was performed for the association of the respective genotypes with (A) fibrosis stage [0-4], (B) steatosis grade [1-3], (C) portal inflammation [0-2], and (D) lobular inflammation [0-2].

Whereas, variants in *HSD17B13* and in *MARC1* were associated with less advanced histological features (Figure 2 & Supplementary Table 5). rs72613567T>TA in *HSD17B13* was strongly associated with lower grade of portal inflammation (P = 1.5×10^-4^) and rs2642438G>A in *MARC1* was associated with lower grade of steatosis (P = 0.02). Both protective variants showed a trend towards a lower fibrosis stage but no significant association on ordinal regression analysis. Similar results were observed using a dichotomous analysis: *PNPLA3* increased odds for the development of both moderate fibrosis (OR 1.33 (95% CI 1.00 - 1.78)) and advanced fibrosis (OR 1.96 (95% CI 1.29 - 3.00)), while *HSD17B13* and *MARC1* were associated with lower odds for moderate fibrosis (OR 0.66 (95% CI 0.45 - 0.99) or advanced fibrosis (OR 0.53 (95%CI 0.30 - 0.94)) respectively (Supplementary Figure 1 & Supplementary Table 6).

There was no association of rs58542926C>T in *TM6SF2* with any histological trait (Supplementary Tables 5 & 6).

### Liver proteomics implicates variants in *HSD17B13* and *MARC1* in fibrosis and lipid metabolism

In order to understand the effect of these variants on liver function, we performed proteomics on liver biopsy samples from 70 children with NAFLD, who were representative of the overall NAFLD group (Supplementary Table 7). rs72613567T>TA was associated with lower abundance of HSD17B13 liver protein (Figure 3A&B & Supplementary Table 8). Whereas hepatic levels of MARC1 protein did not appear to be affected by rs2642438G>A genotype (Figure 3C & Supplementary Table 8).

**Figure 3.**
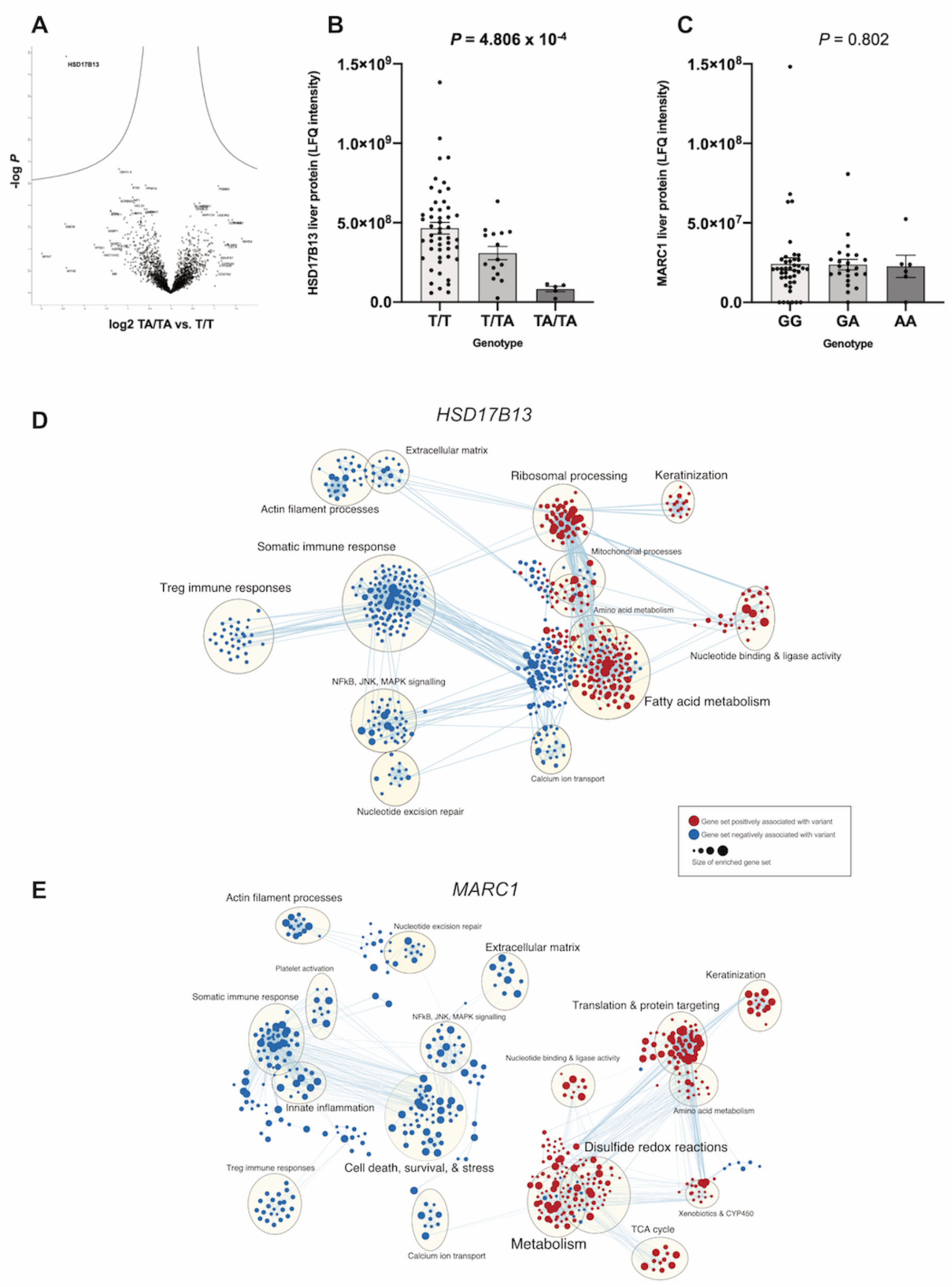
Liver proteomics in children with NAFLD, stratified by rs72613567T>TA in *HSD17B13* & rs2642438G>A in *MARC1* variants. (A) A volcano plot of log2 abundance ratios against the - logio (p-value) of the proteome for the *HSD17B13* TA/TA vs. T/T genotype. (B) Absolute abundance levels (LFQ intensity) stratified by genotype for HSD17B13 liver protein and (C) MARC1 liver protein. Reported *P* values were calculated using Kruskal-Wallis test. (D) Enrichment map of up- (red) or downregulated (blue) pathways for *HSD17B13* genotype as discovered by gene set enrichment analysis using an additive genetic model. (E) Enrichment map of up- (red) or downregulated (blue) pathways for *MARC1* genotype as discovered by gene set enrichment analysis using an additive genetic model.

Gene set enrichment analysis (GSEA) for rs72613567T>TA in *HSD17B13* genotype implicated changes in multiple gene sets, including a strong upregulation of ribosomal activity (e.g. KEGG_RIBOSOME, normalized enrichment score (NES) = 3.0; *Q* = 0) and nonsense-mediated decay (e.g. REACTOME_NONSENSE_MEDIATED_DECAY, NES = 2.74; *Q* = 0), consistent with degradation of mutant *HSD17B13* (Supplementary Table 9). When the enriched gene sets were mapped for similarity (Figure 3D) several trends could be observed: there was a strong signature of downregulation of pathways associated with immune response (e.g. HALLMARK_INTERFERON_GAMMA_RESPONSE, NES = -2.4, *Q* = 0). Multiple metabolic pathways appeared to be perturbed including upregulation of fatty acid processing (e.g. KEGG_FATTY_ACID_METABOLISM, NES = 2.82; *Q* = 0). There was also downregulation of pathways and proteins associated with extracellular matrix formation (e.g. GO_COLLAGEN_CONTAINING_EXTRACELLULAR_MATRIX, NES = -1.9; *Q* = 0.047).

GSEA for rs2642438G>A in *MARC1* similarly found a strong signature for downregulation of extracellular matrix and collagen-related pathways (e.g. GO_COLLAGEN_TRIMER, NES: -2.46, Q = 0.010 and GO_EXTRACELLULAR_MATRIX_STRUCTURAL_CONSTITUENT, NES = -2.35; *Q* = 0.018) as well as gene sets related to the innate immune response (Figure 3E & Supplementary Table 9). There was a strong enrichment in gene sets associated with mitochondrial metabolism, drug detoxification, sphingolipid metabolism, and redox reactions (e.g. GO_SPHINGOLIPID_METABOLIC_PROCESS, NES = 2.15, *Q* = 0.05 and GO_OXIDOREDUCTASE_ACTIVITY, NES = 1.9, *Q* = 0.16).

GSEA for both protective variants demonstrated an upregulation of retinol metabolism (NES = 2.45; *Q* = 4.5×10^-5^ for *HSD17B13* and NES = 2.14; *Q* = 0.03 for *MARC1*).

### *In silico* analysis of MARC1 p.Ala165Thr indicates loss of stability

rs72613567T>TA in *HSD17B13* falls at a splice-site and our proteomics data suggests this results in reduced expression through nonsense mediated decay. However, the effect of rs2642438G>A (coding for p.Ala165Thr) on *MARC1* is less clear, as our proteomics results did not show any change in expression of MARC1 with genotype.

*MARC1* position 165 lies within the cytoplasm (with positions 2-20 within the mitochondrial matrix) and is part of the MOSC (MOCO sulfurase C-terminal) domain. The crystalline structure of *MARC1* has been resolved to 1.78Å and shows that alanine-165 forms part of an alpha-helix on the external surface of the enzyme (Figure 4A&B). Alanine-165 is highly conserved across mammals (Figure 4C), though the Zebrafish isoform of MARC1 has a different structure in this region. Using *in silico* saturation mutagenesis, we observed that Alanine-165 is considered to have a substantial beneficial effect on the protein, compared to other predicted missense variants in *MARC1* (Figure 4D). Whilst position 165 is not within any of the predicted active sites of *MARC1*, two prediction tools classified the p.Ala165Thr variant as disease causing, suggesting that it would cause loss of the alpha-helix and alter the metal binding ability of *MARC1* (Supplementary Table 10). Consistent with this, the p.Ala165Thr variant is also predicted to affect the overall stability of the protein.

**Figure 4.**
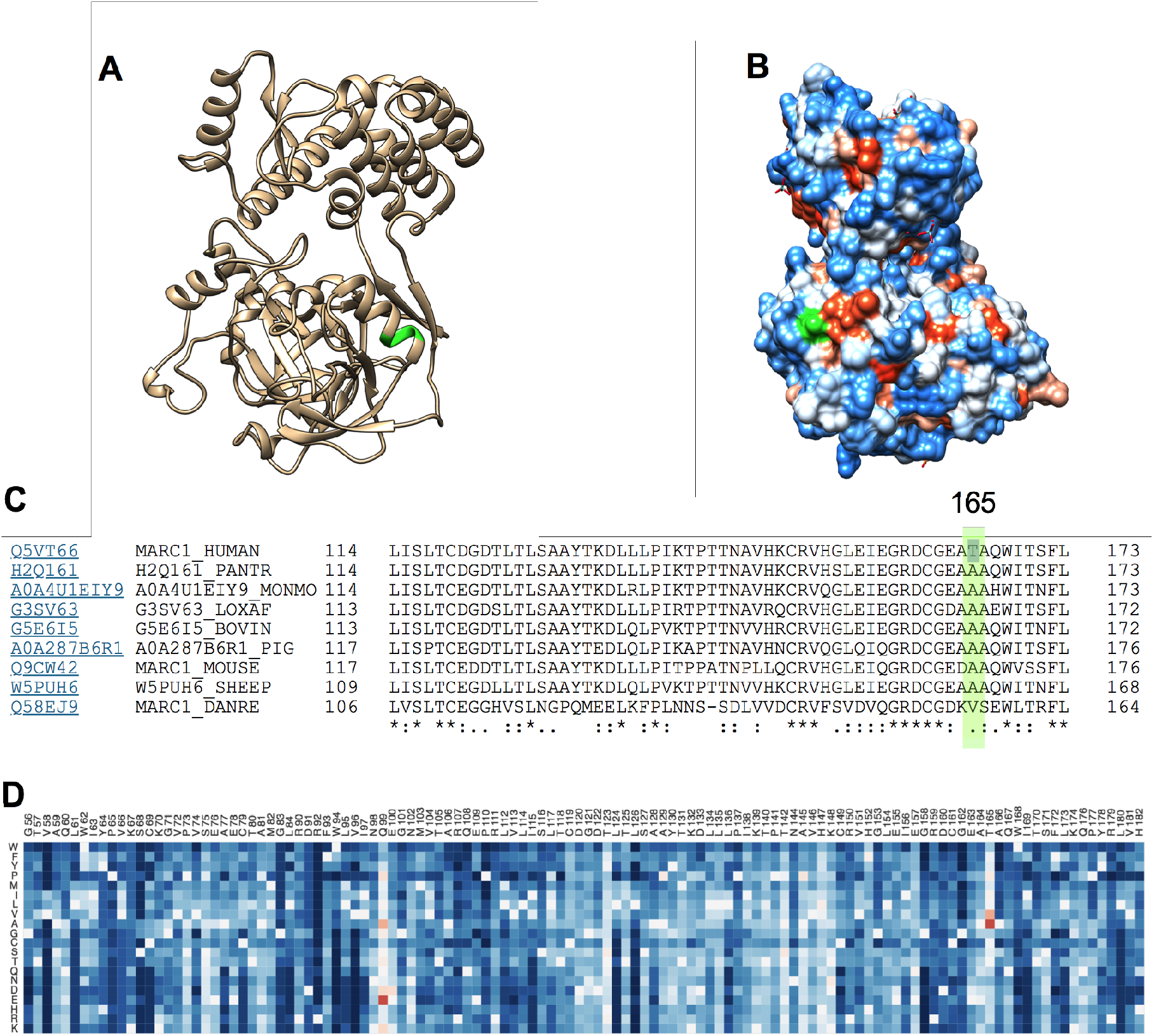
*In silico* prediction of the effects of p.Ala165Thr on *MARC1*. Alanine-165 (green) forms part of an alpha helix (A) and forms part of the external surface of *MARC1* (B). This position is highly conserved in mammals using alignment of protein isoforms (C). Saturation mutagenesis results from EVmutation predicts an increase in protein function (red) when substituting alanine for threonine at position 165 (D).

### Plasma lipidomics shows variants in *HSD17B13 and MARC1* perturb phospholipid metabolism

Given that proteomics data implicates these protective variants in (glycerophospho)-lipid metabolism, we performed untargeted plasma lipidomics in 129 children with biopsy-defined NAFLD and who were representative of the overall NAFLD group (Supplementary Table 11). We tested for associations between variant genotypes and lipid species using linear regression, adjusted for age and sex.

rs72613567T>TA in *HSD17B13* was associated with changes in phosphatidylcholines (PC) and fatty acids (FA) derived from phospholipids (Figure 5A-C). This variant showed a trend towards positive associations with unsaturated FA and negative associations with polyunsaturated FA (Figure 5C). The opposite trend was observed with phosphatidylethanolamines (Supplementary Figure 2). rs2642438G>A in *MARC1* was negatively associated with PC (Figure 5D), again unaffected by chain length (Figure 5E & Supplementary Figure 3), and with phosphatidylinositols (PI, Figure 5F).

**Figure 5.**
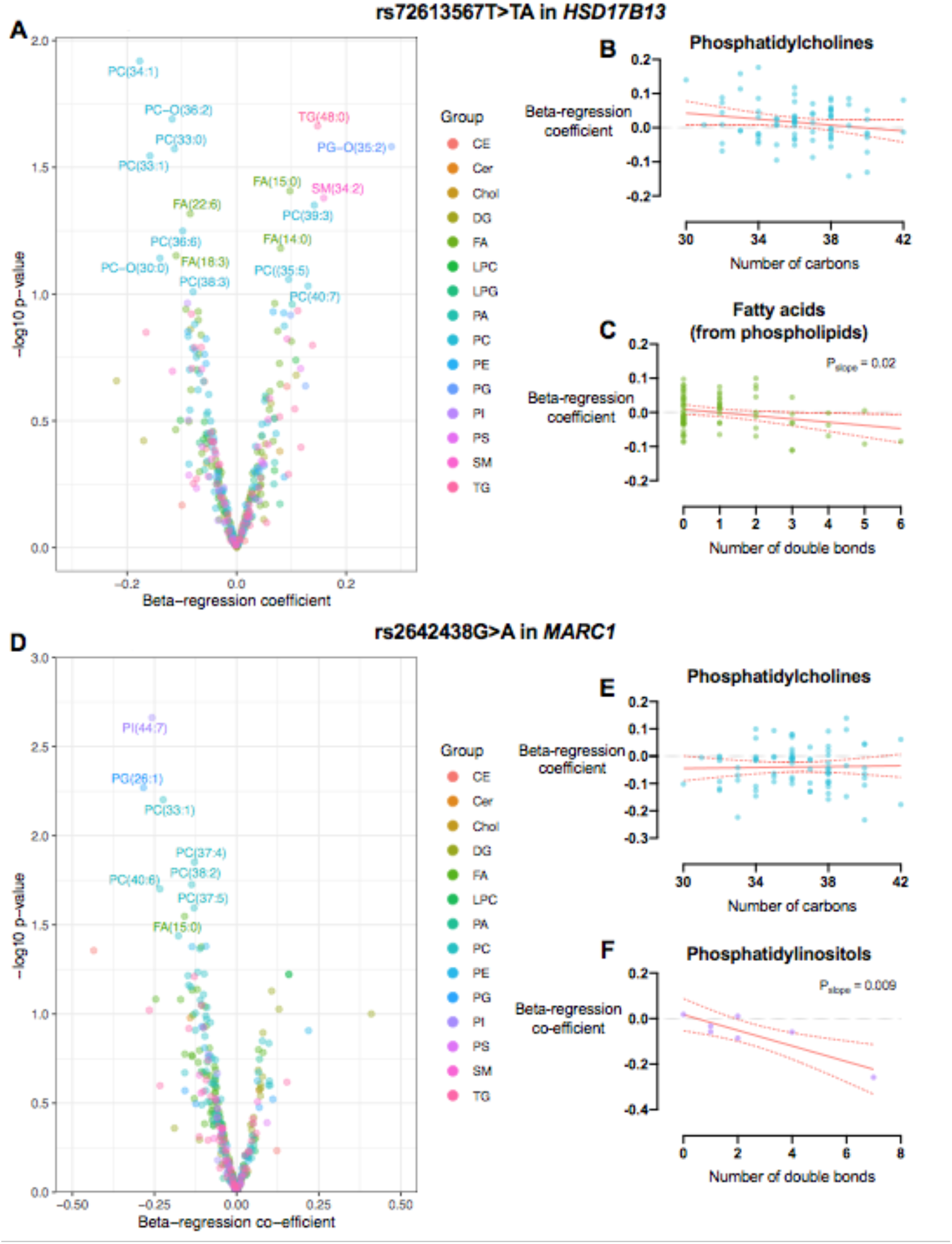
Plasma lipid species associated with rs72613567T>TA in *HSD17B13* and rs2642438G>A in *MARC1*. Volcano plots (A & D) demonstrating the association (as beta-regression coefficient) between lipid species and variants, where beta-regression coefficient was calculated by linear regression between genotype (coding T/T=0, T/TA=1, TA/TA=2) and logarithmically-transformed lipid abundance, adjusted for age and sex. rs72613567T>TA in *HSD17B13* was negatively associated with a higher number of double bonds in fatty acids (FA, B). Whilst rs2642438G>A in *MARC1* was negatively associated with a higher number of double bonds in phosphatidylinositols (PI). Simple linear regression with 95% confidence intervals are shown in red. Data from 129 children with biopsy-confirmed NAFLD. CE, cholesterol esters; Cer, ceramides; Chol, cholesterol; DG, diglycerides; LPC, /yso-phosphatidylcholines; PA, phosphatidic acids; PC, phosphatidylcholines; PE, phosphatidylethanolamines; PG, phosphatidylglycerols; PS, phosphatidylserines; SM, sphingomyelin; TG, triglycerides.

The harmful variant rs738409C>G in *PNPLA3* was also associated with changes in PC, though the direction was influenced by the number of double bonds and carbon chain length (Supplementary Figure 4).

These associations did not appear to be reflective of changes in fibrosis stage, steatosis, or inflammatory activity (Supplementary Figure 5). Fibrosis stage was negatively associated with PC chain length and number of double bonds, whilst NAFLD Activity Score was negatively associated with triglyceride chain length.

### Discussion

In this study we have provided histological validation of protective *MARC1* and *HSD17B13* variants in children with NAFLD. Both variants were associated with perturbed phospholipid metabolism and downregulation of fibrotic pathways. Unlike rs72613567T>TA in *HSD17B13*, hepatic expression of *MARC1* is not affected by p.Ala165Thr, suggesting a mechanism of reduced function.

Both rs72613567T>TA in *HSD17B13^12^* and rs2642438G>A in *MARC1^13^* were originally identified as GWAS-significant risk-reducing loci for liver disease in adults, most recently as risk-reducing variants for all-cause cirrhosis (as well as NAFLD cirrhosis). *HSD17B13* has subsequently been validated in multiple cohorts of adults^6,14,15,29,57-59^ though the histological features associated with this variant in *MARC1* has not been described in children until now^18^. We therefore selected these two variants to study in children, in addition to the well-established risk-increasing locus rs738409C>G in *PNPLA3* and rs58542926C>T in *TM6SF2*.

The splice variant rs72613567T>TA in *HSD17B13* has been consistently associated with lower grade of lobular inflammation, NASH, and stage of fibrosis in adults^12^, though without any difference in severity of histological steatosis^14,29^. In children we observed a strong negative association with grade of peri-portal inflammation with no effect on lobular inflammation. Pediatric NASH is often characterized by a ‘zone 1’ predominant distribution of steatosis and inflammation, particularly in younger children^23^. Whilst it has been speculated that the peri-portal inflammation of pediatric NASH ‘transitions’ to lobular inflammation (and ballooning) of adult NASH, it is challenging to prove. Here, we have observed a genetic variant that has a specific association with peri-portal inflammation in children and with lobular inflammation in adults. We believe this provides further evidence to support the notion that pediatric NASH shares similar genetics to adult NASH, despite having a different histological patterns.

Several groups have previously demonstrated that the splice variant rs72613567T>TA in *HSD17B13* is associated with reduced expression of the enzyme^12,14,29^, which we have replicated using proteomics. Our pathway analysis also suggested an increase in nonsense-mediated decay, similar to the expression profiling data from Sookoian *et a/^29^*. The function of this enzyme is not exactly clear but has been recently implicated in phospholipid metabolism^14^. Our proteomic and lipidomic data is generally concordant with reduced expression of *HSD17B13* causing perturbation of glycerophospholipid metabolism. Though we did not observe precisely the same pattern of altered lipid species as reported by Luukkonen *et al*.^14^, we used plasma, rather than hepatic tissue, for lipid profiling.

The *MARC1* variant discovered by Emdin *et al*. was associated with lower odds of all-cause cirrhosis, diagnosis of fatty liver, and lower liver fat on CT^13^. Consistent with this, in children with NAFLD we found a negative association with severity of steatosis, however we did not replicate the finding of lower odds of NAFLD diagnosis. Whilst there was only a negative trend in fibrosis stage our proteomics data gave a signal of downregulation of fibrotic pathways. One explanation for these findings is that this variant primarily reduces hepatic fat and reduced fibrosis (i.e. cirrhosis in adults) is a secondary effect. Data from other variants associated with liver fat does suggest, via a Mendelian randomization method, that degree of steatosis is causally associated with fibrosis stage^60^. However from our, and others’ observations, the lack of correlation between steatosis and fibrosis stage with rs72613567T>TA in *HSD17B13* suggests that the two histological features can be dissociated. Further mechanistic investigation will be needed to understand the link between these variants, steatosis, and fibrosis.

The precise role of *MARC1* in hepatic metabolism is unknown. We found this enzyme to be expressed at similar levels across rs2642438G>A genotypes, even though the variant (p.Ala165Thr) would be predicted to have a destabilizing effect. *MARC1* is a molybdenum-dependent enzyme that reduces N-oxygenated molecules^17,44^. We found that alanine 165 is highly conserved and threonine-165 may disrupt the alpha-helix and its ability to bind molybdenum. This does appear to affect lipid metabolism as it was associated with reduced abundance of specific plasma phospholipids. Though precisely how this achieved will require further characterization.

Our proteomics showed a consistent trend of increased retinol metabolism associated with *HSD17B13* and *MARC1* variants, whilst it was reduced with the *PNPLA3* variant, as reported previously^27^. Whilst it is possible that a specific common mechanism underlies this, it may also be a secondary observation. Mechanistic work does implicate *PNPLA3^61^* and *HSD17B13* in retinol metabolism^15^ but it is not known for *MARC1*. These results could also be accounted for by activation of hepatic stellate cells^62^ in the context of more advanced NAFLD, with an accompanying down-regulation of their retinol metabolism. More generally it is unclear whether disordered retinol metabolism is causal in severity of NAFLD.

The strengths of this study include a comparatively large number of histologically characterised children and use of unbiased lipidomics and proteomics work to give insights into variant function. Also, replication of well-established associations with rs738409C>G in *PNPLA3* provides further confidence in our findings. Studying paediatric subjects with NAFLD reduces the risk of interaction with factors attributable to adult multimorbidity or substance toxicity.

In this study we were unable to account for genetic ancestry in analyses due to use of genotyping individual variants. Also, we did not observe an association between rs58542926C>T in *TM6SF2* and NAFLD severity, which may have been due to relatively few T-allele carriers. In addition, we may have had reduced power for case-control analyses by use of ultrasound and aminotransferases (rather than more sensitive techniques) for exclusion of steatosis. Therefore, some children with mild steatosis may have been assigned to the control group. Finally, comparatively few children with advanced NAFLD may have meant reduced power to identify further histological associations.

### Conclusion

rs72613567T>TA in *HSD17B13* and rs2642438G>A in *MARC1* are protective against severity of pediatric NAFLD, suggesting shared genetic effects between adults and children. Proteomic and lipidomic data implicate the effect of these two variants with perturbation of phospholipid metabolism in hepatic fibrosis.

## Data Availability

Full summary statistics available in Data Supplement.

## Acknowledgements

The authors would like to thank the help of expert pathology review of liver biopsies from Rita De Vito (Rome) and the contribution of the late Valerio Nobili to this project.

## Grant support

CAH, DM and SW are supported by the German Systems Biology Program “LiSyM” (grant no. 31L0057 and 31L0058) sponsored by the German Federal Ministry of Education and Research (BMBF). JPM is supported by a Wellcome Trust fellowship (216329/Z/19/Z), a European Society for Paediatric Research (ESPR) Young Investigator Award, and a Children’s Liver Disease Foundation Grant. EU-PNAFLD Registry is supported by a European Association for Study of the Liver (EASL) Registry Grant and by the Newcastle NIHR Biomedical Research Centre/Newcastle University using a database infrastructure developed by the EU EPoS consortium. AA is supported by grant from Italian Ministry of Health (5×1000). SF was supported by funding form BBSRC (BB/M027252/1, original proposal written by AK). BK is supported by grants from Van den Broek Lohman Foundation, Virtutis Opus Foundation and For Wishdom Foundation. QMA in a Newcastle NIHR Biomedical Research Centre investigator and a member of the EU H2020 EPoS (Elucidating Pathways of Steatohepatitis) and EU IMI2 LITMUS (Liver Investigation: Testing Marker Utility in Steatohepatitis) consortia funded under grant agreements 634413 and 777377 respectively. No study sponsors had any role in the study design, or data collection, analysis, and interpretation.

**Abbreviations**
CI: confidence intervals;
EU-PNAFLD: European Paediatric NAFLD Registry;
FA: fatty acids;
GSEA: gene set enrichment analysis;
HOMA-IR: homeostatic model assessment of insulin resistance;
HSD17B13: hydroxysteroid 17 beta-13;
MARC1: mitochondrial amidoxime reducing component 1;
NAFLD: non-alcoholic fatty liver disease;
NASH-CRN: Nonalcoholic Steatohepatitis Clinical Research Network;
NES: normalized enrichment score;
OR: odds ratio;
PC: phosphatidylcholines;
PI: phosphatidylinositols PNPLA3, patatin-like phospholipase domain-containing protein 3;
TM6SF2: transmembrane 6 superfamily 2;

## Author contributions

Study concept and design (CAH, JPM, BGK, AV, AA); acquisition of data (CAH, AA, AC, LGD, SF, BJ, KK, DAK, BGK, SGS, IvM, AV, JH, DM, JPM); analysis and interpretation of data (CAH, JPM, BGK, AV, AA, DM, QMA); drafting of the manuscript (CAH, JPM); critical revision of the manuscript for important intellectual content (CAH, AA, AC, LGD, SF, BJ, KK, DAK, AK, BGK, SGS, IvM, AV, JH, DM, JPM, QMA); statistical analysis (CAH, SF, SGS, DM, JPM); obtained funding (CAH, BGK, AV, AA, AK, JPM); administrative, technical, or material support (AC, KK, BJ); study supervision (CAH, JPM).

